# On the reliability of model-based predictions in the context of the current COVID epidemic event: impact of outbreak peak phase and data paucity

**DOI:** 10.1101/2020.04.24.20078485

**Authors:** Jean Daunizeau, Rosalyn Moran, Jérémie Mattout, Karl Friston

**Affiliations:** Institut du Cerveau et de la Moelle épinière, INSERM UMRS 1127, Paris, France; Department of Neuroimaging, Institute of Psychiatry, Psychology & Neuroscience, King’s College London, London, SE5 8AF, UK; Lyon Neuroscience Research Center, CRNL; INSERM, U1028; CNRS, UMR5292; Brain Dynamics and Cognition Team, Lyon, F-69000, France; The Wellcome Centre for Human Neuroimaging, University College London, UK

## Abstract

The pandemic spread of the COVID-19 virus has, as of 20^th^ of April 2020, reached most countries of the world. In an effort to design informed public health policies, many modelling studies have been performed to predict crucial outcomes of interest, including ICU solicitation, cumulated death counts, etc… The corresponding data analyses however, mostly rely on restricted (openly available) data sources, which typically include daily death rates and confirmed COVID cases time series. In addition, many of these predictions are derived before the peak of the outbreak has been observed yet (as is still currently the case for many countries). In this work, we show that peak phase and data paucity have a substantial impact on the reliability of model predictions. Although we focus on a recent model of the COVID pandemics, our conclusions most likely apply to most existing models, which are variants of the so-called “Susceptible-Infected-Removed” or SIR framework. Our results highlight the need for performing systematic reliability evaluations for all models that currently inform public health policies. They also motivate a plea for gathering and opening richer and more reliable data time series (e.g., ICU occupancy, negative test rates, social distancing commitment reports, etc).

## 1. Introduction

As with the situation almost exactly one hundred years ago -with the Spanish flu-the pandemic spread of the COVID-19 virus has, as of 20^th^ of April 2020, reached most countries around the world (Johnson and Mueller, 2002). The entire scientific community is now addressing the many issues that the virus poses, with an unprecedented collaborative spirit. One of these issues is of primary importance for guiding national and international decision makers: namely, predicting the health requirements and outcomes of the current epidemiologic event (Siedner et al., 2020; Wang, 2020). Over the past two months, about a thousand modelling papers have been deposited on preprint servers such as ArXiv or MedRXiv (Nature, 2020). This, if anything, demonstrates how fast and efficiently the scientific community can be set in motion towards a common goal. However, it is now apparent that models have not reached consensus, e.g., when it comes to predicting the population levels acquired immunity within the next months (Moran et al., 2020). This is unfortunate, since this - and related predictions- are critical for designing suppression or mitigation strategies that aim at limiting the human cost of the current pandemic (Canabarro et al., 2020; James et al., 2020; Rodriguez et al., 2020).

Most models that attempt to furnish such predictions are variants of the SIR framework (Kermack et al., 1927). In brief, these models assume that the population is divided into, e.g., ‘Susceptible’, ‘Infected’, and ‘Removed^1^’ compartments, through which individuals transit at a pace that is characteristic of the time course of the infection and associated socio-medical measures. Under mild assumptions, all SIR models predict that the population dynamics will eventually reach so-called ‘disease-free equilibria’, which signal the end of the epidemic outbreak by exhaustion of the ‘susceptible’ compartment. This means that the signature of an epidemic lies in the transient dynamics of observable health reports such as confirmed case numbers and mortalities. These models have proven very useful in predicting, e.g., the prevalence or the duration of an epidemic. When properly adjusted to observable epidemiologic data, they also can serve to predict the impact of candidate health policies such as vaccination or social distancing (Ganem et al., 2020; Jeria et al., 2020; Kissler et al., 2020; Moghadas et al., 2020). Given the past success of these models, it may then come as a surprise that, despite relying on the same data sources, these models do not make the same predictions. In this note, we argue that this lack of consensus arises *because* modellers use the same dataset^2^, which comprises cumulative counts of death and positive COVID testing. The critical question here is: can these data sufficiently constrain estimates of SIR cycles? If not, then subtle variants of SIR models may make dramatically different predictions, despite showing almost no difference in terms of the fit accuracy on the available data so far (Salomon, 2020). Also, model predictions may depend sensitively upon the current phase of the epidemic event: more precisely, whether the outbreak peak has been reached or not (Lin et al., 2020). This is because the ramping phase of the epidemic transient may not evince all the processes that are relevant for estimating unknown model parameters (and hence making reliable model-based predictions).

In this note, we assess the impact of outbreak peak phase and data paucity on the reliability of predictions derived from SIR-type models. In particular, we evaluate the prediction accuracy of a recent SIR-type model that follows from augmenting the set of data to be explained (in particular, we focus on ICU occupancy and negative testing rates^3^, in addition to positive test results and death rates records), depending on whether the outbreak has already been observed or not.

## 2. Methods

### a. The DCM-covid model

In what follows, we will focus on a specific SIR model, namely: the so-called dynamic causal model of COVID pandemics or DCM-covid (Friston et al., 2020; Moran et al., 2020). In brief, the model considers four interacting factors describing location, infection status, test status and clinical status, respectively. Within each factor people may probabilistically transition among four distinct states. Given a set of 21 model parameters (see below), the model describes the temporal dynamics of the marginal probabilities of belonging to each state within each factor. The location factor describes if an individual is at home, at work^4^, in an intensive care unit (ICU) or in the morgue. The infection status is the closest to native SIR models, and includes susceptible, infected, contagious or immune states. Note that, at this point, the model assumes that the immune state is absorbing, i.e. people cannot get the disease twice. The clinical status factor comprises asymptomatic, symptomatic, acute respiratory distress syndrome (ARDS) or deceased. Finally, the diagnostic status captures the fact that a given individual can be untested, waiting for the results of a test, or declared either positive or negative. Model transitions amongst states are controlled by rate constants (inverse time constants) and probability constants (e.g., the probability of dying when in ICU). The ensuing set of state probabilities can then be related to some specific observable epidemiologic outcomes, such as the number of deceased people per day or the number of people newly infected who have been tested positive. Figure 1 below summarizes the causal structure implicit in conditional transition probabilities. We refer the reader to Friston et al. (Friston et al., 2020) for a complete mathematical description of the model.

**Figure 1:**
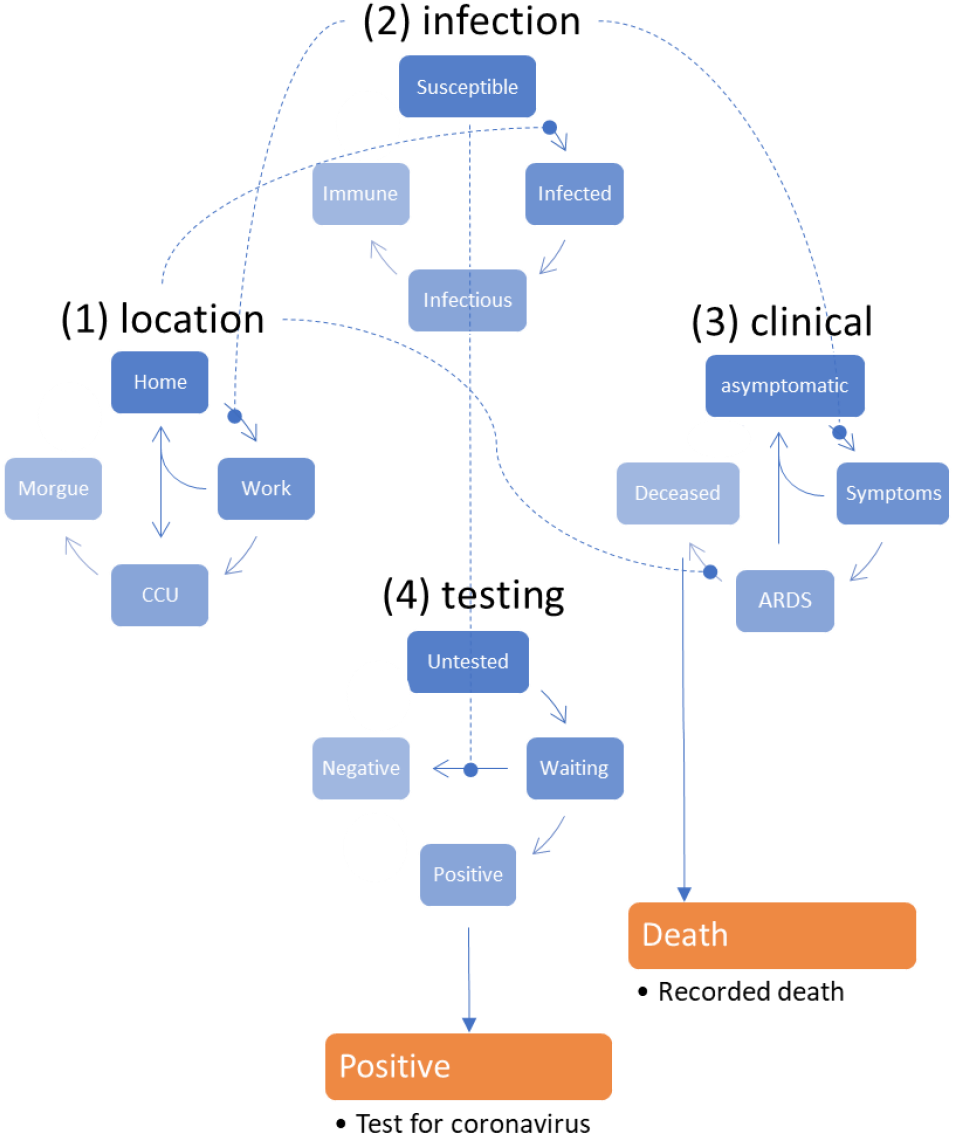
Causal structure of the DCM-COVID model (adapted from Friston et al. 2020). In brief, this compartmental model generates timeseries data based on a mean field approximation to ensemble or population dynamics. The implicit probability distributions are over four latent factors, each with four levels or states (see main text). In particular, this model assumes that (i) there is a progression from a state of susceptibility to immunity, through a period of (pre-contagious) infection to an infectious (contagious) status, (ii) there is a progression from asymptomatic to ARDS, where people with ARDS can either recover to an asymptomatic state or not. With this setup, one can be in one of four places, with any infectious status, expressing symptoms or not and having test results or not. Note that—in this construction—it is possible to be infected and yet be asymptomatic. Crucially, the transitions within any factor depend upon the marginal distribution of other factors. For example, the probability of becoming infected, given that one is susceptible to infection, depends upon whether one is at home or at work. Similarly, the probability of developing symptoms depends upon whether one is infected or not. The probability of being testing negative depends upon whether one is susceptible (or immune) to infection, and so on. Finally, to complete the circular dependency, the probability of leaving home to go to work depends upon the number of infected people in the population, mediated by social distancing. At any point in time, the probability of being in any combination of the four states determines what would be observed at the population level. For example, the occupancy of the deceased level of the clinical factor determines the current number of people who have recorded deaths. Similarly, the occupancy of the positive level of the testing factor determines the expected number of positive cases reported. From these expectations, the expected number of new cases per day can be generated. A more detailed description of the generative model can be found in Friston et al (Friston et al., 2020).

Parameter estimation and model comparison relies on a variational Bayesian approximation scheme (Daunizeau, 2018; Friston et al., 2007) which is adopted in established computational neuroscience toolboxes (Ashburner, 2012). In this particular work, we have chosen to implement the DCM-covid model from scratch, and make it available for another open academic model-based data analysis toolbox (Daunizeau et al., 2014). We did this to provide software validation for subsequent data analyses performed with the DCM-covid model.

In addition to semi-informed prior distributions, model inversion –in the current implementation-places hard constraints on parameters to ensure that they stay within admissible ranges. More precisely, rate constants and probability constants are derived by passing unbounded parameters through an exponential mapping and sigmoid mapping, respectively^5^. Table 1 below recapitulates the unknown model parameters, in terms of its prior mean and associated hard constraint.

**Table 1:** Summary of priors for DCM-covid model parameters. Note that prior variances were fixed to 1 for all unbounded model parameters

| Number | Parameter | Mean | Description |
| --- | --- | --- | --- |
| 1 | $\theta_n = \exp(\mathcal{G}_n)$ | 1 | Number of initial cases |
| 2 | $\theta_N = s(\mathcal{G}_N) \times N_{country}$ | 1/2 | Initial number of susceptible people ($N_{country}$ is the corresponding country) |

<sup>5</sup> Astute readers will notice a few minor changes from the original model inversion scheme proposed by Friston et al. (Friston et al., 2020). In particular, the hard sigmoid constraint on transition probability parameters ensures that these cannot be greater than one.

|  |  |  |  |
| --- | --- | --- | --- |
|  |  |  | population size<sup>6</sup>, in millions) |
| **Location** |  |  |  |
| **4** | $\theta_{out} = s(\mathcal{G}_{out})$ | 1/3 | Prob(*work* \| *home*): prob of going out |
| **5** | $\theta_{sde} = \exp(\mathcal{G}_{sde})$ | 1 | Social distancing exponent |
| **6** | $\theta_{cap} = s(\mathcal{G}_{cap})$ | 128/1000<br>00 | ICU capacity threshold (per capita) |
| **Infection** |  |  |  |
| **7** | $\theta_{Rin} = \exp(\mathcal{G}_{Rin})$ | 3 | Effective number of contacts: home |
| **8** | $\theta_{Rou} = \exp(\mathcal{G}_{Rou})$ | 48 | Effective number of contacts: work |
| **9** | $\theta_{trn} = s(\mathcal{G}_{trn})$ | 1/4 | Prob(*contagion* \| *contact*) |
| **10** | $\theta_{inf} = \exp(-\frac{1}{\tau_{inf}})$ | $\tau_{inf} = 5$ | Infected (pre-contagious) period (days) |
| **11** | $\theta_{con} = \exp(-\frac{1}{\tau_{con}})$ | $\tau_{con} = 3$ | Infectious (contagious) period (days) |
| **Clinical** |  |  |  |
| **12** | $\theta_{dev} = s(\mathcal{G}_{dev})$ | 1/3 | Prob(*symptoms* \| *infected*) |
| **13** | $\theta_{sev} = s(\mathcal{G}_{sev})$ | 1/100 | Prob(*ARDS* \| *symptomatic*) |
| **14** | $\theta_{sym} = \exp(-\frac{1}{\tau_{sym}})$ | $\tau_{sym} = 5$ | symptomatic period (days) |
| **15** | $\theta_{rds} = \exp(-\frac{1}{\tau_{rds}})$ | $\tau_{rds} = 12$ | acute RDS period (days) |
| **16** | $\theta_{fat} = s(\mathcal{G}_{fat})$ | 1/3 | Prob(*fatality* \| *CCU*) |
| **17** | $\theta_{sur} = s(\mathcal{G}_{sur})$ | 1/16 | Prob(*survival* \| *home*) |
| **Testing** |  |  |  |
| **18** | $\theta_{tfr} = s(\mathcal{G}_{tfr})$ | 500/1000<br>00 | Threshold: testing capacity (per capita) |
| **19** | $\theta_{sen} = s(\mathcal{G}_{sen})$ | 1/100 | Rate: testing capacity (%) |
| **20** | $\theta_{del} = \exp(-\frac{1}{\tau_{del}})$ | $\tau_{del} = 2$ | Delay: testing capacity (days) |
| **21** | $\theta_{tes} = s(\mathcal{G}_{tes})$ | 1/8 | Relative Prob(*tested* \| *uninfected*) |

We will pay particular attention to estimates of *θ*_*N*_* i.e. the initial number of susceptible people among the country’s population. This is because this parameter eventually controls the quantitative predictions regarding specific outcomes of interest, e.g. acquired immunity ratio at the end of this epidemiologic event. Note that this type of prediction is critical, because it determines the likelihood of multiple rebounds (i.e. waves) of the COVID pandemic are when or if confinement measures are relaxed (Moran et al., 2020).

### b. Observable outcomes

Most modelling studies to date actually rely on daily WHO^7^ or ECDC^8^ data reports, which gathers cumulative death, positive test and remission counts across countries. These dataset are made openly available as part of a global collaborative effort to fight against the COVID pandemic (see: https://github.com/CSSEGISandData/COVID-19). However, remission rates are typically considered unreliable, as is evident from established international worldwide data repositories that prefer to report consolidated death and confirmed positive test counts only (see: https://github.com/owid). This effectively reduces the available data to the death and positive test counts, on which most model predictions rely, including outcomes of interest that are only indirectly informed by these data (e.g., acquired population immunity at the end of the current epidemic outbreak). However, a few governmental agencies have recently made an effort to assemble and make openly available richer datasets, including, e.g., ICU occupancy and confirmed negative test counts (see, e.g., for France: https://www.data.gouv.fr/fr/datasets/). This is particularly relevant in this modelling context, because recent SIR models comprise multiple compartments that capture modern health care practices that are only partially observable (see, e.g.: https://ecosys.versailles-grignon.inra.fr/SpatialAgronomy/covid19/).

As with most modelling studies currently performed on the COVID pandemic, previous applications of the DCM-covid model only fitted daily death (hereafter *o*^(1)^) and positive test (*o*^(2)^) counts. However, the structure of the model and its associated inversion scheme makes it very easy to augment the generated outcome data with remission^9^ (*o*^(3)^), ICU occupancy (*o*^(4)^), and negative test (*o*^(5)^) rates:

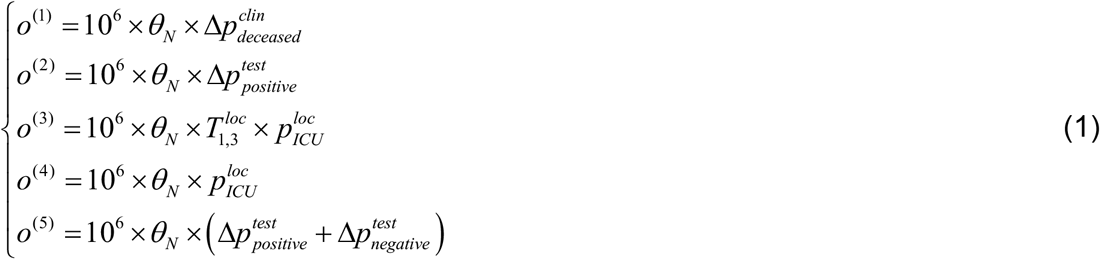

Where 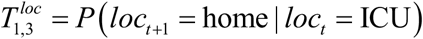 is the daily probability of transiting from ICU to home given that the previous clinical status was ARDS and Δ*p* ≜ *p*_*now*_ − *p*_*yesterday*_ is the daily change in the corresponding marginal probability (with a slight abuse of notation)^10^. The particular form Equation 1 takes for confirmed cases and death counts, derives from the fact that model inversion focuses on fitting newly (daily) counts, as opposed to cumulative counts (with the exception of remission rates and ICU occupancy).

One can see from Equation 1 that observable outcomes provide only partial information regarding internal (latent or hidden) model states. At this point, we note that, in principle, nothing prevents such SIR models to be fitted to other types of observable data. However, at the time of writing this manuscript, no other data type was reliably and regularly accessible. We therefore focus on these five observable outcomes.

### c. Numerical assessment of prediction accuracy

At the time of writing, most western European countries (including, e.g., UK, Germany, France, Italy, Spain, Ireland and Switzerland) have either passed the peak of the pandemic event or are about to pass it. Available data typically start on January the 20^th^, yielding approximately a hundred daily data points up until the current time. But the transient dynamics of the COVID pandemic will not approach disease-free equilibria states for a further 6 to 8 months. This means that one has to predict what will happen over the next 200 days, given what has happened during the past 100 days. As we will see, the accuracy of the ensuing model predictions actually depends upon how far the country is with respect to the peak of its epidemic outbreak, in terms of the death rate. The accuracy of these predictions will also depend upon what type of data is actually provided to the model inversion.

We thus simulated 1000 datasets with varying phases of the peak, which could emerge either before or after the first (available) 100 data samples. We did this by randomly sampling the parameter set around the estimated parameters for the French gouv.fr dataset (up to the 12^th^ of April, see below), which gathers the five outcomes of interest. For each parameter set, we simulated the DCM-covid model over a duration of 300 days. This yielded realistic variations of epidemic outbreak dynamics. Figure 2 below depicts a representative subset of simulated time series, as well as Monte-Carlo histograms of simulated cumulative death counts, initial number of susceptible people and death rate time-to-peaks.

**Figure 2:**
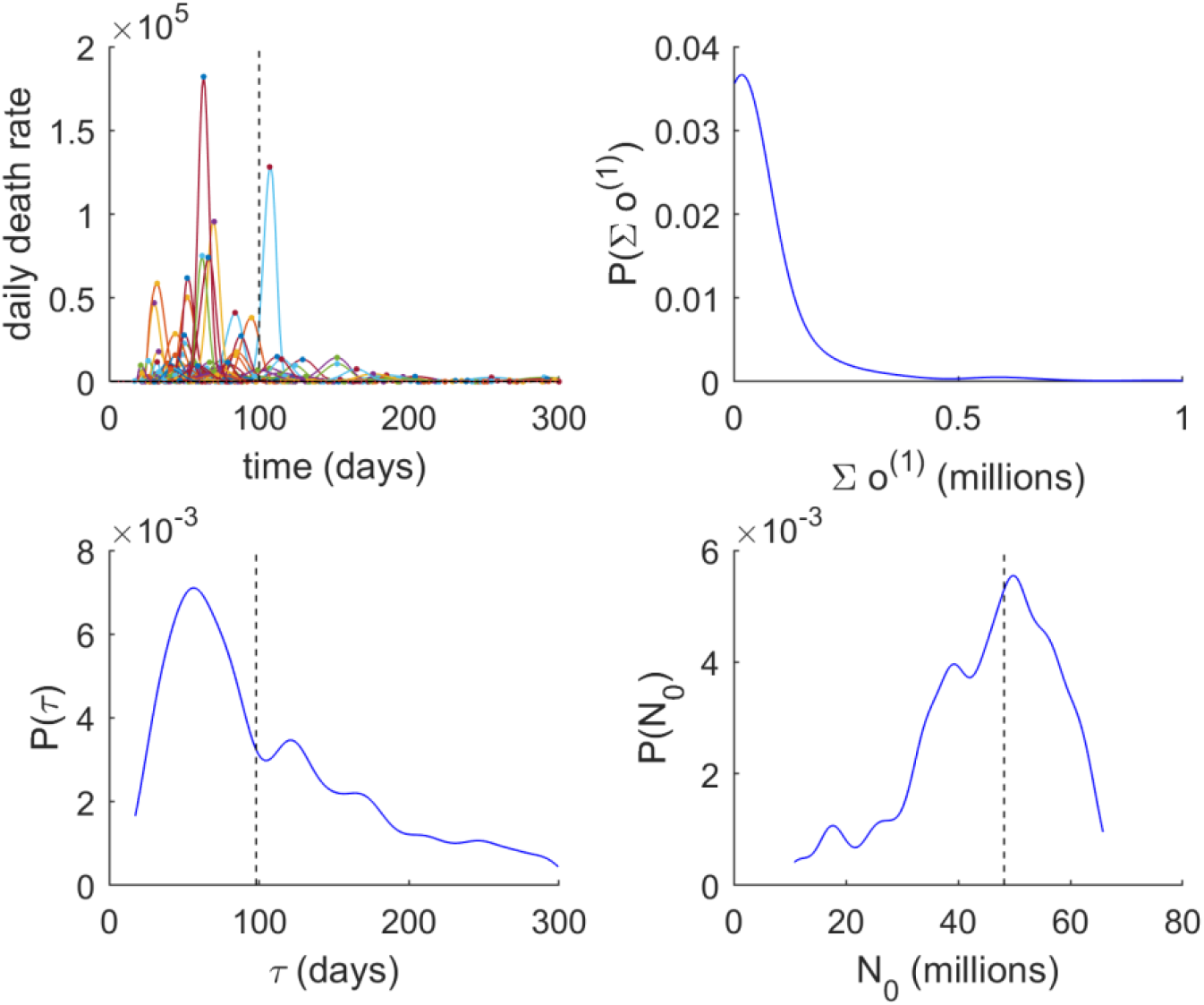
Summary of simulated outcomes of interest. These simulations were drawn from small variations around the model parameters estimated from the full French data available on the 12^th^ of April. Upper-left panel: simulated daily death rate dynamics (y-axis) are plotted against time (x-axis, in days). Dots depict time-to-peaks. Upper-right panel: the distribution of simulated cumulated death counts. Lower-left panel: the distribution of simulated time-to-peaks. Lower-right panel: the distribution of simulated initial number of susceptible people (in millions).

One can see that all generated outcome dynamics exhibit a simple transient, eventually reaching the disease-free equilibrium state (where 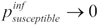). In addition, one can see that simulations are collectively reminiscent of the variability observed across different countries.

Each simulated data time series was then truncated up to the 100^th^ day, and then fitted using the DCM-covid VB inversion scheme. We considered two inversion variants:

- VB0: the full dataset (comprising the five observable outcomes) is provided (up to the 100^th^ day) to the VB inversion scheme.
- VB1: remission, ICU and negative test time series are omitted (this is the typical situation for most modelling studies so far).

For each simulated dataset, we thus obtained two sets of estimated parameters, one from each VB inversion schemes. We derive the ensuing predictions by simulating the model for the remaining 200 days, given each of those estimated parameter sets.

- Peak date estimation error, which we define as follows:

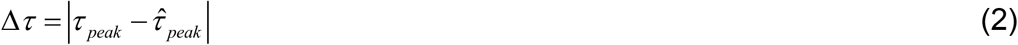

where *τ*_*peak*_and 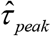 are the simulated and estimated outbreak peak, respectively.
- Cumulated death count prediction error:

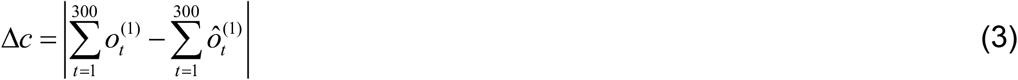

where 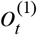 and 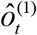 are the simulated and estimated daily deceased rates, respectively.
- Initial susceptible ratio estimation error:

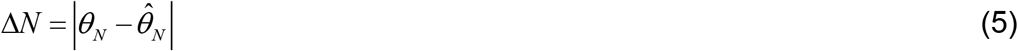

where *θ*_*N*_ and 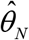 are the simulated and estimated daily deceased rates, respectively.
- Maximum ICU occupancy:

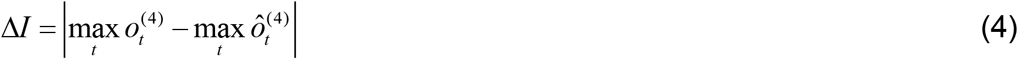

where 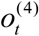 and 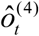 are the simulated and estimated daily ICU occupancy, respectively.

We then analysed the impact of the time-to-peak (*τ*_*peak*_) and data paucity (cf. the two VB variants) onto the four prediction/estimation error scores above.

## 3. Results

### a. Influence of date-to-peak and dataset availability on prediction/estimation accuracy

Figure 3 below depicts the relationship between simulated and estimated outcomes (more precisely: epidemics’ peak date, cumulative death count, maximum ICU occupancy and initial number of susceptible people).

**Figure 3:**
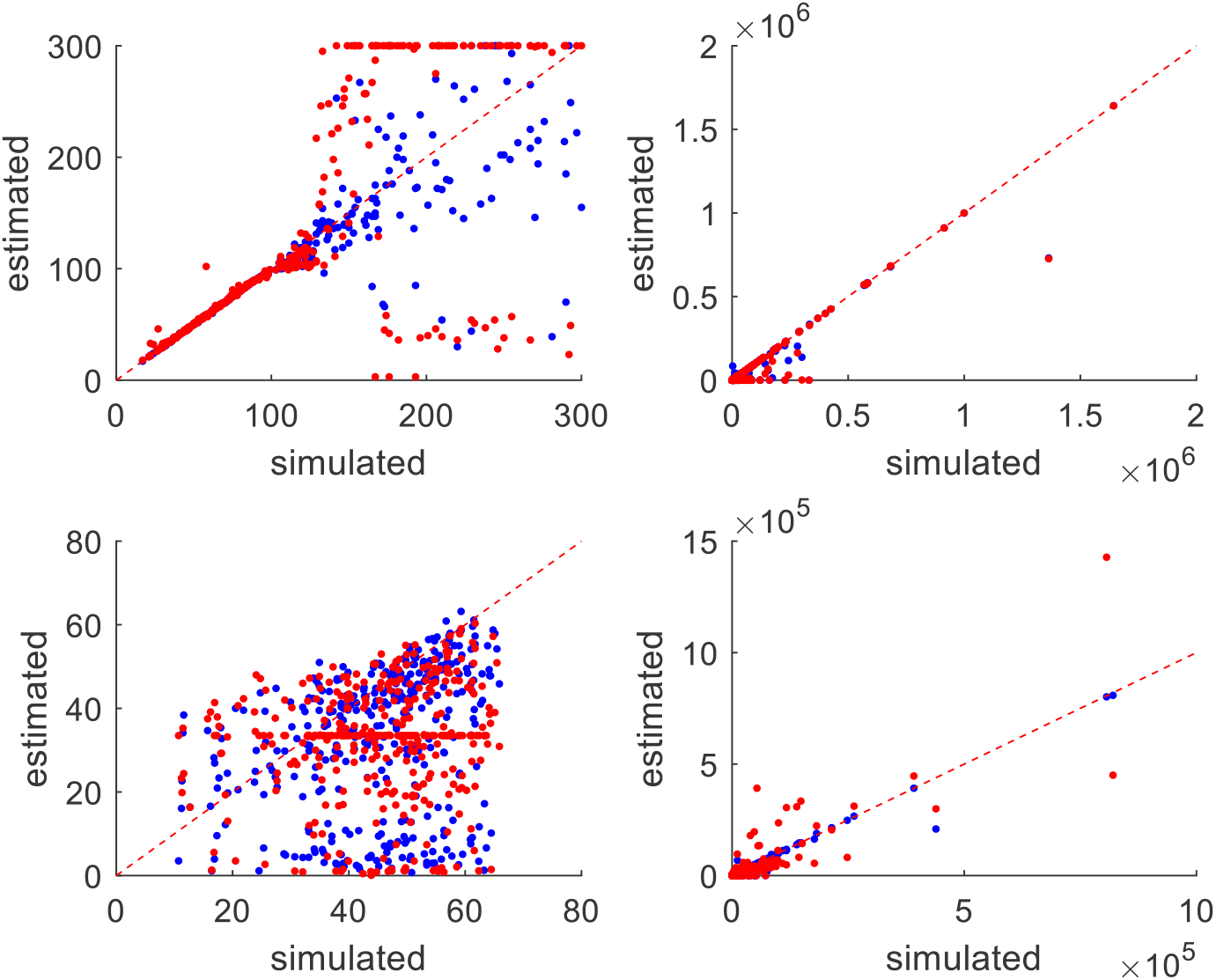
Relationship between simulated and estimated/predicted outcomes of interest. Upper-left panel: estimated time-to-peaks (y-axis, in days) are plotted against simulated time to peaks (x-axis, in days), for both VB0 (blue, augmented data) and VB1 (red, positive cases and death rates only) data analyses. Each dot corresponds to a simulated data time series, and the red dotted line shows the identity mapping (perfect estimation/prediction). Upper-right panel: cumulated death counts,. Lower-left panel: initial number of susceptible people,. Lower-right panel: maximum ICU occupancy.

One can see that this relationship is highly variable, i.e. estimation/prediction errors are clearly non-negligible. Importantly, these errors are not due to model underfitting, since the percentage of explained variance in fitted data (i.e. data up to 100 days) is always greater than 95% (VB0: mean R^2^=99%, VB1: mean R^2^=99%). Therefore, estimation/prediction errors are due to non-identifiability. The structure of these errors is most apparent for time-to-peak (cf. upper-left panel in Figure 3). When the simulated time-to-peak *τ* _*peak*_ < 100, the correlation between simulated and estimated time-to-peaks in the initial (observed) 100 days is almost perfect. However, this correlation quickly falls as the simulated time-to-peak increases beyond the temporal window of observed data (and more so when fitted data is restricted to daily death counts and positive test rates: VB1). The structure of estimation/prediction errors is less explicit for outcomes of interest. We now evaluate the impact of simulated time-to-peak and data availability on estimation/prediction errors.

First, we split the simulations according to whether the simulated death rate peak arose before the last observed sample (*τ* < 100, “early peak”) or after (*τ* > 100, “late peak”). This enabled us to ask whether the prediction/estimation errors above were higher for late than for early peaks. Figure 4 below summarizes the simulation results, in terms of the influence of time-to-peak (early versus late peak) and dataset availability (VB0 versus VB1) onto prediction/estimation accuracy.

**Figure 4:**
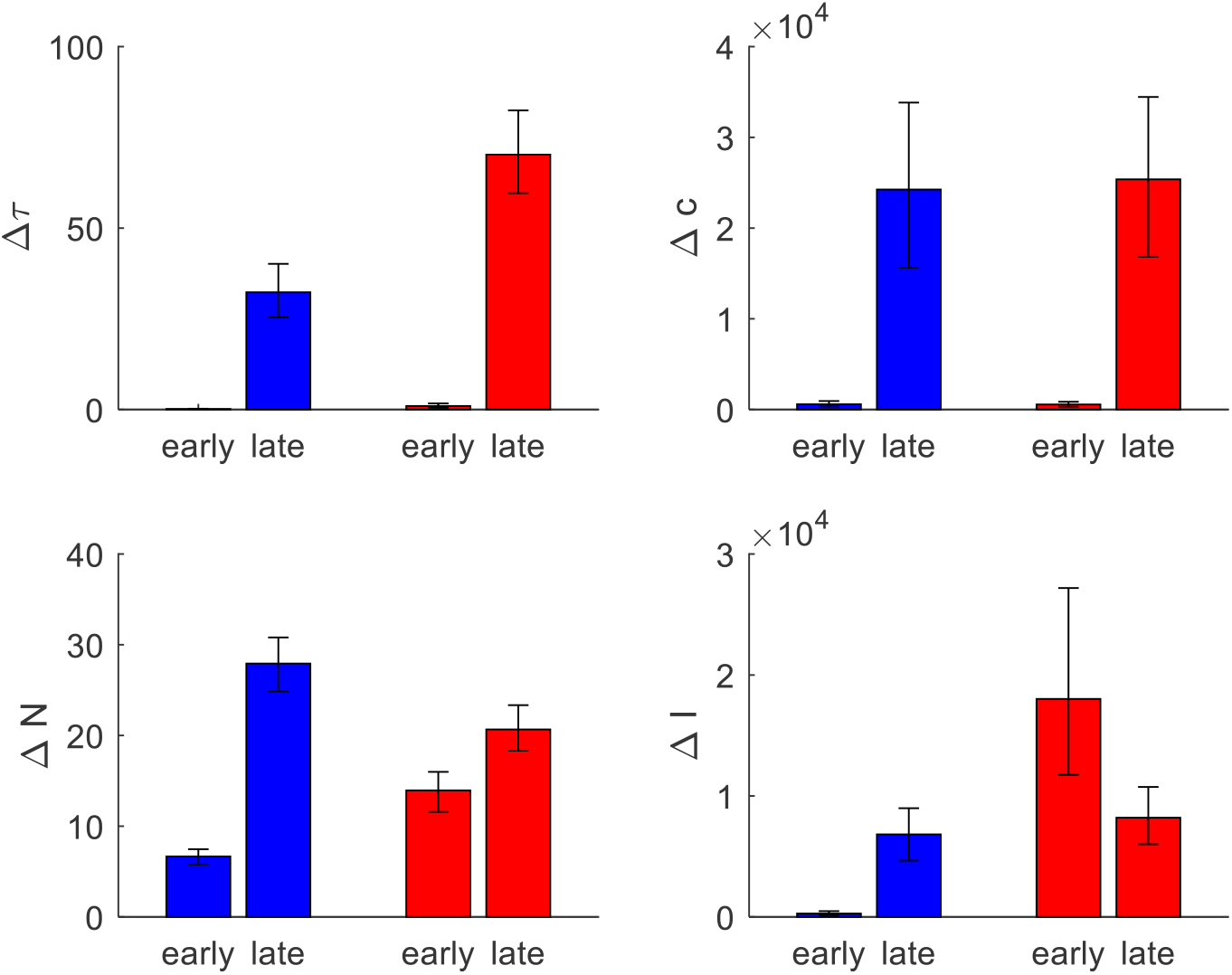
Impact of time-to-peak and data availability on estimation/prediction accuracy. Upper-left panel: Average peak date prediction error Δ*τ* (y-axis, in days) is shown for early and late peaks, for both VB0 (blue) and VB1 (red) data analyses. Errorbars denote 95% bootstrapped confidence intervals on the mean. Upper-right panel: cumulated death counts error Δ*c*, same format as upper-left panel. Lower-left panel: initial number of susceptible people error Δ*N*, same format as upper-left panel. Lower-right panel: maximum ICU occupancy, same format as upper-left panel.

To begin with, note the typical error magnitude for the four outcomes of interest: time-to-peak error is of the order of 20 days, cumulative death count error is about 10,000 people, the error on the initial number of susceptible people is 10 millions, and the maximum ICU occupancy error is around 10,000. These errors are beyond acceptable limits, for most practical applications to public health policies. But, as we will see, error magnitudes depend sensitively upon time-to-peak and data paucity.

One can see a clear influence of the time-to-peak onto all prediction/estimation error measures. In brief, it seems that both prediction and estimation are much less accurate when they are performed before the death rate peak has been observed (late peak). The influence of the data availability (VB0 vs VB1) is less clear, though it seems that, depending on the outcome of interest, restricting the dataset may decrease accuracy for both early and late peaks.

To confirm these observations, we performed a simple 2×2 ANOVA on each of the four error scores. First, the main effect of time-to-peak is significant for all error types except for ICU occupancy (Δ*τ* : p<10^−4^, Δ*c* : p<10^−4^, Δ*I* : p=0.84, Δ*N* : p<10^−4^). Second, there is a significant main effect of data availability in two out of four error types (Δ*τ* : p<10^−4^, Δ*c* : p=0.44, Δ*I* : p<10^−4^, Δ*N* : p=0.59). Note that the two-way interaction between time-to-peak and data availability is statistically significant for all error types except for cumulative death count (Δ*τ* : p<10^−4^, Δ*c* : p=0.88, Δ*I* : p<10^−4^, Δ*N* : p<10^−4^). This does not create an interpretational issue for the associated main effect of time-to-peak however, since there is no crossover interaction (except for ICU occupancy). We then performed post-hoc tests.

It transpires that, when the peak can be observed in the fitted data (early peaks), VB0 accuracy is significantly higher than VB1 accuracy for both ICU occupancy (p<10^−4^) and initial number of susceptible people (p<10^−4^). In contrast, when the peak is yet to manifest (late peaks), only the accuracy on time-to-peak estimation is significantly worse for VB1 than for VB0 (p<10^−4^). Interestingly, the ICU occupancy error is highest for VB1 when the peak has already been observed (cf. lower right panel). This counter-intuitive result derives from the fact that default model explanations of death and positive test rates dynamics favour overestimated ICU occupancy (which, for VB1, is not constrained by ICU occupancy data). This will be clearer when analysing the French dataset below.

In summary, the reliability of almost all predicted outcomes is severely impacted when the data analysis is performed before the epidemic peak has been observed. In addition, ignoring data such as ICU occupancy and negative test rates strongly impairs the estimation of the initial number of susceptible people as well as the maximum ICU occupancy, in particular when data is analysed >100 days before the epidemic peak has been observed.

### b. Example: French data

We will now illustrate our analysis of the reliability of the model’s prediction/estimation using a single country’s data. We focus on French data, because governmental agencies provide additional data^11^, which are missing from WHO or ECDC databases (as of today). Note that reported daily death rates are restricted to hospital data, i.e. it does not include those people who do not die in hospitals (c.f. e.g., retirement homes).

We pre-processed the time series to correct data reports from various counting errors (see below). We also padded the governmental data with ECDC data from the 1^st^ of January to the 18^th^ of March (for both daily death and positive test rates) because these dates are not reported in the online available governmental data repositories. This means that there are missing data for both ICU occupancy and total test rates. Figure 5 below shows the effect of data smoothing on the observed data.

**Figure 5:**
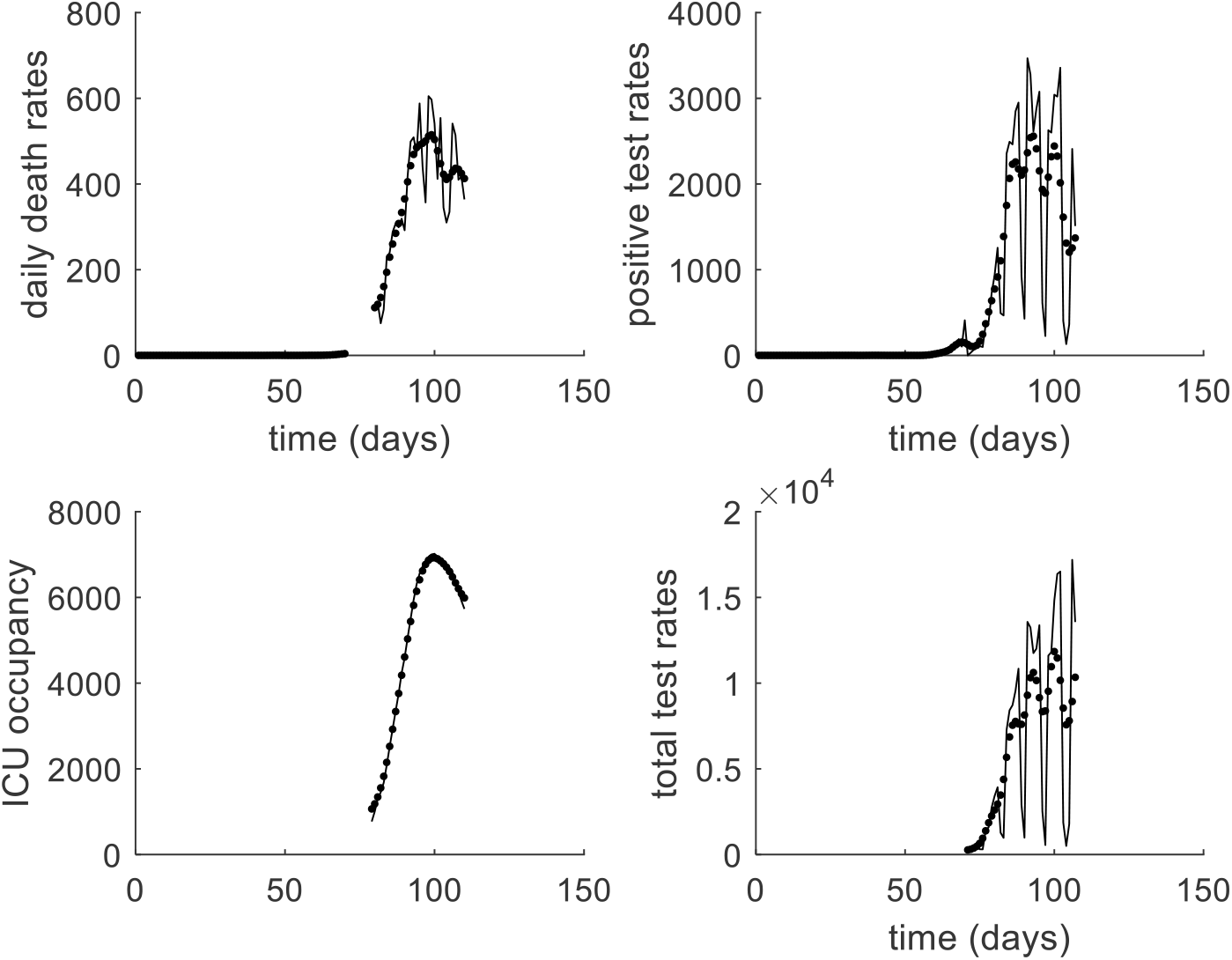
Data pre-processing. Upper-left panel: Reported daily death rate dynamics (y-axis) is plotted against time (x-axis, in weeks), starting on the 1^st^ of January 2020 and ending on the 19^th^ of April 2020. The black line and black dots denote the uncorrected and corrected data, respectively. Upper-right panel: positive test rates, same format as upper-left panel. Lower-left panel: total test rates, same format as upper-left panel. Lower-right panel: ICU occupancy error Δ*I*, same format as upper-left panel.

One can see that the native positive and total test rates exhibit strong periodic dips. Inspection of the corresponding dates show that these dips correspond to data reports made on weekends. Data pre-processing corrects most of these inconsistencies, without impacting on the corresponding cumulated counts (not shown).

We conducted two analyses on these corrected datasets, by either fitting all reported data (VB0) or only daily death and positive test rates (VB1). Figure 6 below summarizes the ensuing data fits and their predicted dynamics 100 days beyond the last reported date.

**Figure 6:**
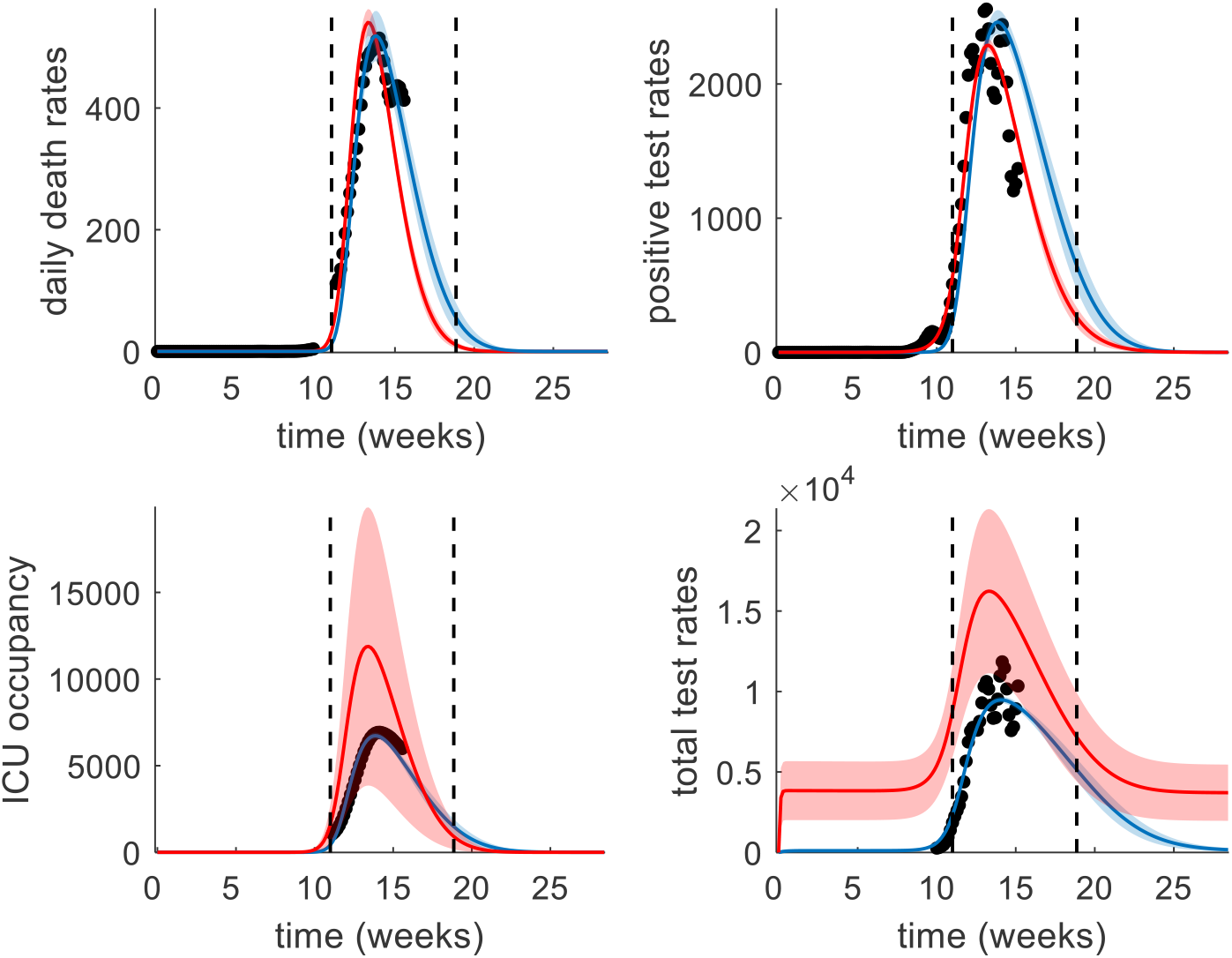
Model fits of available French data: impact of data paucity. Upper-left panel: Reported daily death rate dynamics (y-axis) is plotted against time (x-axis, in weeks), starting on the 1^st^ of January 2020 and ending on the 19^th^ of April 2020. Note: only hospital mortality is reported here. The blue and red traces denote the predicted data when accounting for ICU occupancy and negative test rates and when not accounting for these, respectively. The black dots show the corrected available data. Black dotted lines denotes the start and intended end of the governmental containment measures (17^th^ of March and 11^th^ of May, respectively). Upper-right panel: positive test rates, same format as upper-left panel. Lower-left panel: ICU occupancy. Lower-right panel: total test rates.

Recall that VB0 and VB1 both attempt to account for observed daily death rates and positive test rates. One can see that both succeed in explaining these time series with very high accuracy. In fact, VB1 explains these data better (VB0: R^2^[death rate]=99%, R^2^[positive test rate]=94%, VB1: R^2^[death rate]=98%, R^2^[positive test rate]=99%). However, only VB0 tries to concurrently fit ICU occupancy and negative test rates. Here again, observed time series are very well explained (VB0: R^2^[ICU occupancy]=95%, R^2^[total test rate]=95%). In contrast, VB1’s estimates of these time series are substantially overestimated. This is because VB1 has (unknowingly) overfitted the positive test rates, which has resulted in parameter estimation errors. The situation is quite different for VB0, which had to find parameter estimates that yield a balanced trade-off between all concurrent data reports. This observation recapitulates the simulations results regarding ICU occupancy error (although France is currently lying in between typical early or late peak phases).

Although fit accuracies for common datasets are comparable, VB0 and VB1 make remarkably different predictions. This is illustrated on Figure 7 below.

**Figure 7:**
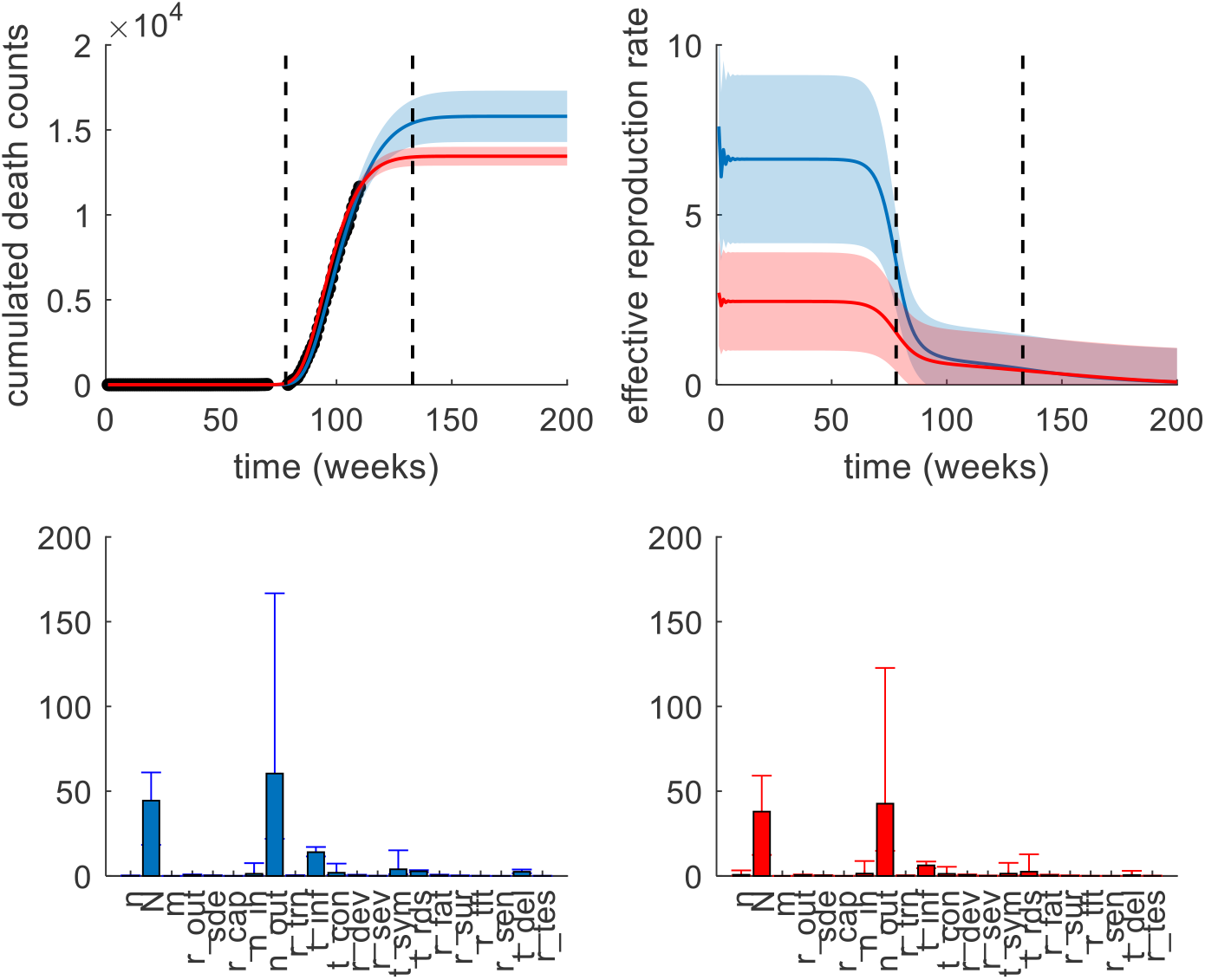
Model predictions/estimations given available French data. Upper-left panel: Reported cumulated death count dynamics (y-axis) is plotted against time (x-axis, in weeks). Same format as Figure 6. Note: only hospital mortality is reported here. Upper-right panel: effective reproduction rate, same format as upper-left panel. Lower-left panel: estimated parameters given all available data (VB0). Errorbars show Bayesian 95% posterior credible intervals. Lower-right panel: estimated parameters, when ignoring ICU occupancy and negative test rates (VB1), same format as upper-left panel.

First, the estimated peak date is 6^th^ of April for VB0, whereas it is 2^nd^ of April for VB1 (this can be seen in Figure 6). Second, the predicted cumulated death counts after the current epidemic outbreak clearly differ. For VB0, predicted cumulated death counts should be 15799 +/- 255 (11635, as of 18-Apr-2020), whereas for VB1, predicted cumulated death counts should be 13450 +/- 124. In brief, ignoring ICU occupancy and negative test rates yield epidemic outbreaks that terminate sooner and are less severe (in terms of casualties).

Third, recall that the model can be used to derive estimates of effective reproduction rates (R0), i.e. the expected number of people who are infected by a COVID-carrier. This summary statistics of the infectiousness of the epidemics varies over time, depending upon the probability that people stay at home or not (this changes the effective number of social contacts), and the probability of being susceptible to the disease. We refer the interested reader to Equation 1.9 in Friston et al (Friston et al., 2020). It turns out that model-based estimates of the effective reproduction rates’ dynamics are clearly higher when accounting for ICU occupancy and negative test rates (cf. upper-right panel on Figure 7). Note that the effective reproduction rate starts to decrease roughly at the date of public lockdown (Tuesday the 17^th^ of March). This is interesting because the model is not informed about this public health event. More precisely, it defines social distancing in terms of the (hidden) behaviour of citizens, without assuming that everyone follows the governmental containment instructions.

Finally, accounting for ICU occupancy and negative test rates produce more uncertain parameter estimates (cf. lower panels on Figure 7). This is most likely because VB1 overfits the observed data, effectively yielding underestimated (overconfident) evaluations of parameter estimation uncertainty.

## 4. Discussion

In this work, we have evaluated the reliability of model-based estimations/predictions for four outcomes of interest in the context of the current COVID pandemics. We have shown that the reliability of these predictions depends sensitively upon whether they are derived before or after the epidemic outbreak peak. In addition, we have shown that data paucity (in particular, ignoring ICU occupancy and negative test rates) can accentuate these prediction errors, even when the outbreak peak has already been observed. This is crucial when estimating the initial number of susceptible people, given that it determines the immunity ratio acquired by the population at the end of the epidemic event (Moran et al., 2020). We have also illustrated the impact of discounting ICU occupancy and negative test rates on French data available to date. This is a timely analysis, since France is, in all likelihood, currently experiencing the peak of the current epidemic outbreak.

The outbreak peak is a significant marker of the rise and fall of distinct transient epidemic dynamics (and its associated public health measures), the late phase of which is crucial to inform parameter estimation. This is reminiscent of what could be observed in DCMs for neural responses, where for instance, the key role of feedback connections between neuronal populations can only be evidenced once the first peak of the electrophysiological evoked response has been observed (Garrido et al., 2007). Here, some key hidden biological processes (and their associated unknown parameters) can only be reliably inferred after the peak of the transient dynamics.

Having said this, the reliability of model-based predictions for countries that have not passed the peak yet (as is still the case now) could in fact be improved by informing the parameter estimation with data from countries where the outbreak peak has already been observed, using, e.g., hierarchical empirical Bayes models (Friston et al., 2020; Kass and Steffey, 1989). At the European level in particular, this speaks to a common effort to gather and share data.

From a statistical perspective, one may not be surprised that prediction/estimation errors decrease when augmenting the fitted data with ICU occupancy and negative test rates. What is remarkable however, is the quantitative difference it makes for e.g., cumulative death counts or effective reproduction rates (see Figures 6 and 7). More remarkable is the interaction of data paucity and timing of predictions with respect to the outbreak peak. More generally, in those particular times where uncertainty is high and decisions have to be made as quickly as possible, it may be particularly important to complement models with quantitative assessments of their reliability and the limits of our predictive approaches.

As a side point, we have not addressed the reliability of the data we have used in our analysis. Daily death counts, for example, are potentially problematic for at least two reasons. First, different data repositories effectively give different numbers, e.g., people deceased in hospitals (as is the case for the French data we have presented here), or in hospitals and retirement homes. Second, they may not account for “normal” seasonal mortality (Goldstein et al., 2012), though this is not the case here (because these hospital death counts are confirmed COVID cases). Testing procedures also have imperfect sensitivity and specificity (Patel et al., 2020), and ICU occupancy actually depend upon heterogeneous clinical criteria (e.g., respiratory support versus reanimation). All these limitations are difficult (though not impossible) to account for, and further challenge even further the reliability of model-based predictions.

Contrary to most papers that focus on model definition and extension, the approach here tackles this assessment which we believe will become more and more important as more alternative models are proposed, to account for, e.g., the influence of lockdown decisions. This applies to the DCM-COVID model we evaluate here, which is currently being refined along these lines. The kind of data that may need to be acquired to inform the ensuing model predictions is an issue of primary practical importance if this or similar models are to guide public health decisions.

Performing this type of analysis for currently available models is beyond the scope of the current work. However, our results highlight the need for evaluating the reliability of model predictions that are currently used by national and international socio-political decision makers. They also motivate the gathering of multiple data time series and making them available to the modelling community. This requirement obviously extends beyond ICU occupancy and negative test rates (Chen et al., 2020; Salomon, 2020). In the near future for instance, data about the number of asymptomatic cases in the population, about how infectious are children or about individual immunity after recovery may prove critical. In order to validate model predictions, particularly those related to infected or clinical status, biological assays of these inferred measures are required. Serological surveys for example are being rolled out to examine community infection rates. In a recent study in the Santa Clara region of California antibodies to SARs-CoV-2 were identified in 1.5% of 3,330 people sampled – with an adjusted population prevalence of 2.4% to 4.26% of the population (Bendavid et al., 2020), with similar rates identified in an analysis of Dutch blood samples in line with model estimates (Moran et al., 2020). Larger ‘serosurveys’ will ultimately be required to more precisely define these measures with large populations being enrolled currently in Germany and by the US National Institute of Health. In addition, reports from centres of recent outbreaks are providing further details that can inform model parameter priors. For example two hospitals in New York City have recently reported a mechanical ventilation requirement for 33.1% of patients admitted for the treatment of Covid-19 (Goyal et al., 2020). The impact of these and other kinds of data on the reliability of model-based predictions could be evaluated with the approach presented here, irrespective of the model used.

## Data Availability

All data are available upon request to the corresponding author.

## Footnotes

1 “Removed” individuals are either cured (immune) or dead.

2 Most modelling studies actually use the Johns Hopkins University Center COVID database, which gathers data from WHO and other national and international health organizations. It produces daily reports of deaths, positive tests and remission cases for most countries in the world (https://coronavirus.jhu.edu/map.html).

3 These kinds of data are made openly available by some governmental organizations (e.g., Santé Publique France).

4 Here, being at “work” essentially means being neither at home, at the hospital or in the morgue (cf., e.g., children at school).

5 Astute readers will notice a few minor changes from the original model inversion scheme proposed by Friston et al. (Friston et al., 2020). In particular, the hard sigmoid constraint on transition probability parameters ensures that these cannot be greater than one.

6 Country population sizes are taken from available online governmental data.

7 World Health Organization (https://www.who.int/).

8 European Centre for Disease Prevention and Control (https://www.ecdc.europa.eu/en).

9 Here, we are restricting remissions to people leaving ICU and returning home.

10 Note that marginal probability 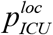 on the third line of Equation 1 is evaluated on the day before, such that 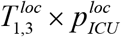 is the current transition rate towards the location status ‘home’.

11 These data are made available online here: https://www.data.gouv.fr/en/datasets/donnees-hospitalieres-relatives-a-lepidemie-de-covid-19/ and here: https://www.data.gouv.fr/en/datasets/donnees-relatives-aux-tests-de-depistage-de-covid-19-realises-en-laboratoire-de-ville/.

